# Prevalence and associated factors of stress among adolescents in rural Bangladesh: A cross-sectional study

**DOI:** 10.1101/2025.08.01.25332676

**Authors:** Md Parvez Shaikh, Rifa Tamanna Mumu, Shadman Sakib Ayan

## Abstract

Adolescence is a critical developmental stage characterized by heightened vulnerability to psychological disorders. However, research on adolescent stress in Bangladesh is limited. This study aims to assess the prevalence and associated factors of stress symptoms among adolescents in rural Bangladesh.

This study is a secondary analysis of a cross-sectional study conducted among 500 adolescents aged 11–17 years in a rural subdistrict of Bangladesh using the Depression, Anxiety, and Stress Scale-21 Items (DASS-21). Chi-square tests and ordinal logistic regression were conducted to identify predictors.

The overall prevalence was 36.2% (mild: 10.6%; 95% CI: 8.2%–13.6%; moderate: 10.8%; 95% CI: 8.4%–13.8%; severe: 9.8%; 95% CI: 7.5%–12.7%; extremely severe: 5.0%; 95% CI: 3.4%–7.3%). Associated factors included older age (AOR: 2.9; 95% CI: 1.7–5.0; p < 0.001), female gender (AOR: 1.8; 95% CI: 1.1–2.9; p = 0.019), being married or in a complex relationship (AOR: 6.1; 95% CI: 1.7–22.5; p = 0.006), high socioeconomic status (AOR: 2.2; 95% CI: 1.2–4.1; p = 0.01), poor parental relationships (AOR: 2.2; 95% CI: 1.2–4.1; p = 0.01), bullying (AOR: 1.9; 95% CI: 1.1–3.5; p = 0.031), peer conflict (AOR: 1.8; 95% CI: 1.1–2.9; p = 0.017), moderate (AOR: 4.9; 95% CI: 1.7–14.3; p = 0.004) and severe educational stress (AOR: 9.1; 95% CI: 2.8–29.9; p < 0.001), and a family history of suicide (AOR: 4.0; 95% CI: 1.6–10.0; p = 0.003).

These findings highlight the urgent need for targeted mental health interventions to reduce stress and promote adolescent psychological well-being.

## Background

Adolescence, commonly defined as the period beginning with puberty and continuing into adulthood, is characterized by significant neural, behavioral, and biological transformations [1]. According to the World Health Organization, adolescence is a period between 10 and 20 years [2]. It is also the developmental stage during which many mental health disorders first emerge, even if they are not diagnosed until later in life [3]. Adolescent mental health has become an increasingly prominent public health priority [4, 5], with stress recognized as one of the most prevalent yet frequently underestimated concerns [6, 7]. Stress in adolescents refers to psychological and physiological responses to demands that surpass their coping capacity, often triggered by academic pressure, interpersonal relationships, and family dynamics [8, 9].

The global prevalence of stress is 35.1%, and stress is more prevalent in high-income countries (HICs) [10]. In Southeast Asian countries, the prevalence of psychological distress among 12-to 15-year-olds is 11.0%, which is higher among females (11.8%) than among males (10.1%) [11]. The prevalence of stress among 13–19-year-old South Indian adolescents is 29.89% [12], and stress is more common in rural areas [13]. Rural adolescents are particularly vulnerable to unique stressors, including academic pressure, economic hardship, romantic relationships, and strained interactions with peers, teachers, and family members [14].

Globally, adolescents encounter a diverse array of stressors [15] during critical phases of physical [16], emotional [17], and cognitive development [18, 19]. in low- and middle-income countries (LMICs) such as Bangladesh [20], these challenges are exacerbated by social stigma, limited mental health services, and inadequate public health infrastructure [21]. The population of Bangladesh was 163.05 million in 2017, and the total mental health expenditure per person was 0.1 USD. Approximately 0.5% of the government budget was allocated for mental health. The number of psychiatrists and psychologists per million population was 1.7 and 3.5, respectively [22], which indicates that the topic of mental health is neglected in Bangladesh. A study indicated that approximately 65% of adolescents in the capital city of Bangladesh suffer from moderate stress, whereas 9% experience high-stress symptoms [23]. Although mental health awareness is gradually increasing in urban Bangladesh, adolescents in rural areas remain substantially underserved [24]. While a few studies have examined the prevalence and determinants of stress symptoms in urban contexts [23, 25], no comparable research has been conducted in rural areas. This study aims to identify the prevalence of stress symptoms and examine their associated factors among rural Bangladeshi adolescents through a cross-sectional survey. Addressing this knowledge gap is critical for designing targeted, evidence-based interventions to improve adolescent psychological well-being in these underserved settings.

## Method

### Study design

This study is a secondary analysis of a cross-sectional study that was conducted from April 15 to May 14, 2024, in three secondary schools located in Lohagara, a rural subdistrict of Narail in southern Bangladesh.

### Study participants

The original study included the target population of adolescents aged 11–17 years residing in Lohagara. The study sample included students enrolled in one government secondary school and two non-government secondary schools within the subdistrict.

### Sample size and sampling technique

Based on a reported 65% prevalence of moderate stress symptoms among adolescents in Bangladesh [23], with a 95% confidence level, and a 5% margin of error, The sample size was calculated 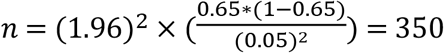

A total of 500 data points were collected via a cluster sampling technique to minimize selection bias. In Bangladesh, secondary schools typically consist of six academic levels—classes **VI** to **X**—along with a class for secondary school certificate (SSC) examination candidates. Each class is divided into three sections, each comprising approximately 50 to 60 students. For this study, four sections from the government school and three from each nongovernment school were randomly selected via simple random sampling.

Nonresident Lohagara students with cognitive impairments, who were unwilling to participate, and who failed to sign the informed written consent form, were excluded from the study. Participants taking medications for mental health disorders were also excluded to minimize confounding, as psychotropic medications may influence mood and behavior.

### Data collection and measurement tools

The DASS-21 scale was used to evaluate the presence and severity of depression, anxiety, and stress. The validity and reliability of this questionnaire were established in previous studies [26-28]. This questionnaire comprises 21 items divided into three subscales (seven items each). Responses were recorded on a 4-point scale ranging from 0 (“Did not apply to me at all”) to 3 (“Applied to me very much or most of the time”). Subscale scores were summed and multiplied by two to obtain the final scores for each domain [29-31]. The participants who identified with symptoms of stress had their legal guardians informed.

Another structured questionnaire was used to collect sociodemographic, behavioral, socioenvironmental, institutional, and family-related information. We adopted the socioecological model (SEM) to guide our conceptual framework. This model suggests that mental health outcomes, such as stress, are influenced by a broader range of factors in individual, interpersonal, institutional, community, and societal domains [32]. All the questionnaires were translated into Bengali to ensure clarity and effective communication with the participants.

### Data management and analysis

Data quality was ensured through consistency verification and double-entry checks. Only patients with complete outcome data were included. Missing data for independent variables were minimal (<5%) and were addressed via listwise deletion. Statistical analyses were performed via STATA version 17, with chi-square tests employed to assess associations and determine p-values. Variables with p-values < 0.05 were selected for ordinal logistic regression to adjust for potential confounders. Odds ratios and their 95% confidence intervals (CIs) indicate the strength of associations. A p-value < 0.05 was considered statistically significant.

### Ethical Considerations

This study utilized previously collected IRB-approved data. Ethical approval for the original data collection was obtained from the Institutional Ethics Committee of North South University (2024/OR-NSU/IRB/0201) before data collection.

Permission was obtained from the respective school authorities. Informed written consent was obtained from the legal guardians of all the participating students, and informed written assent was obtained from each student. The participants were assured of the confidentiality of the information provided and were informed that all the data would be collected anonymously and used solely for research purposes. The present study is a secondary analysis of the dataset.

## Result

### Sociodemographic characteristics

A total of 500 responses were collected, with a 100% response rate. The majority of participants (64.6%, n = 323) were aged between 14 and 17 years. A total of 51.6% (n = 175) of the samples were female. Approximately 44.9% (n = 210) belong to large families, and 12.6% (n = 62) reported that their parents did not own a house. Socioeconomic status was determined by monthly family income, homeownership, and parental occupation. Each indicator reflecting disadvantage was assigned a score (low family income=1, no homeownership=1, unskilled or unemployed parents=1). The total score ranged from 0 to 3. Scores of 0 to 1 indicate high socioeconomic status, 2 indicates medium socioeconomic status, and 3 indicates low socioeconomic status. On this basis, nearly half of the participants (47.6%, n = 238) were classified as having medium socioeconomic status. Most participants (85.4%, n = 426) identified as Muslim. A small proportion (4.3%, n = 20) were engaged in part-time employment in addition to their studies. The majority (78.4%, n = 388) lived with both parents, whereas 8.0% (n = 40) reported having stepparents. Additionally, 3.2% (n = 16) were married or in a complex relationship. **Table 1**.

**Table 1.**
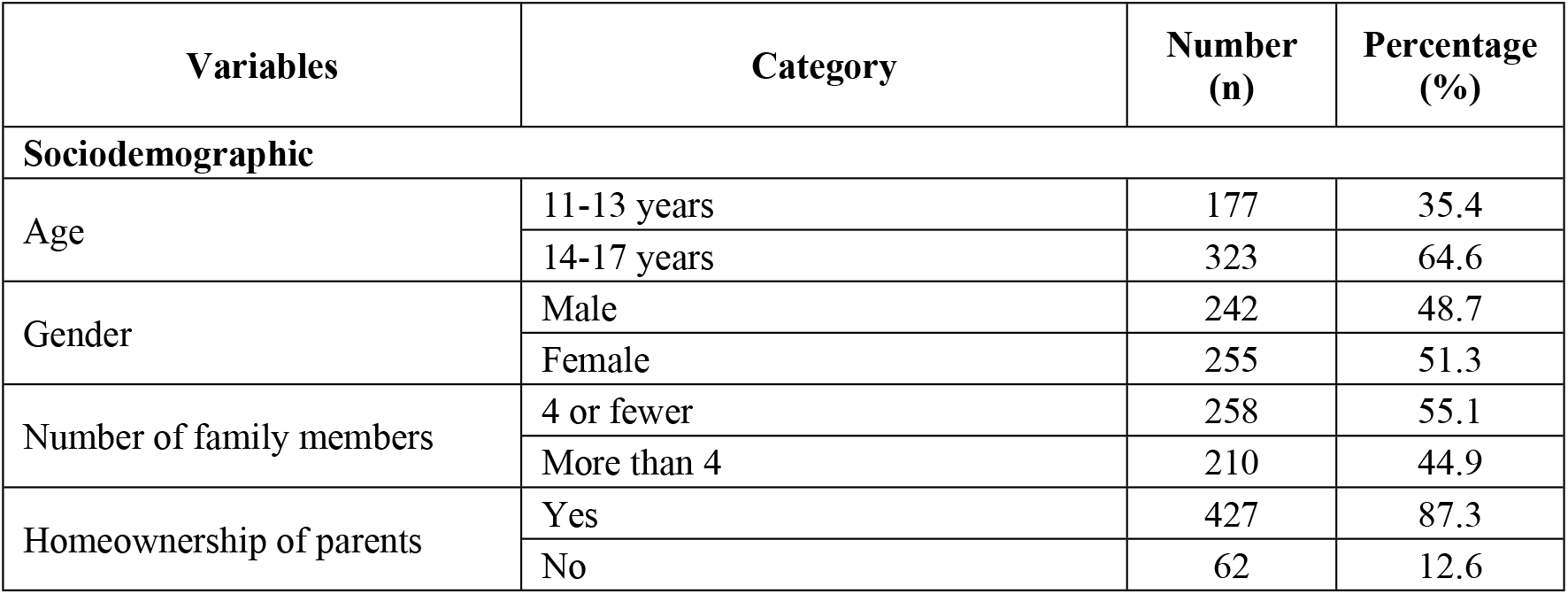
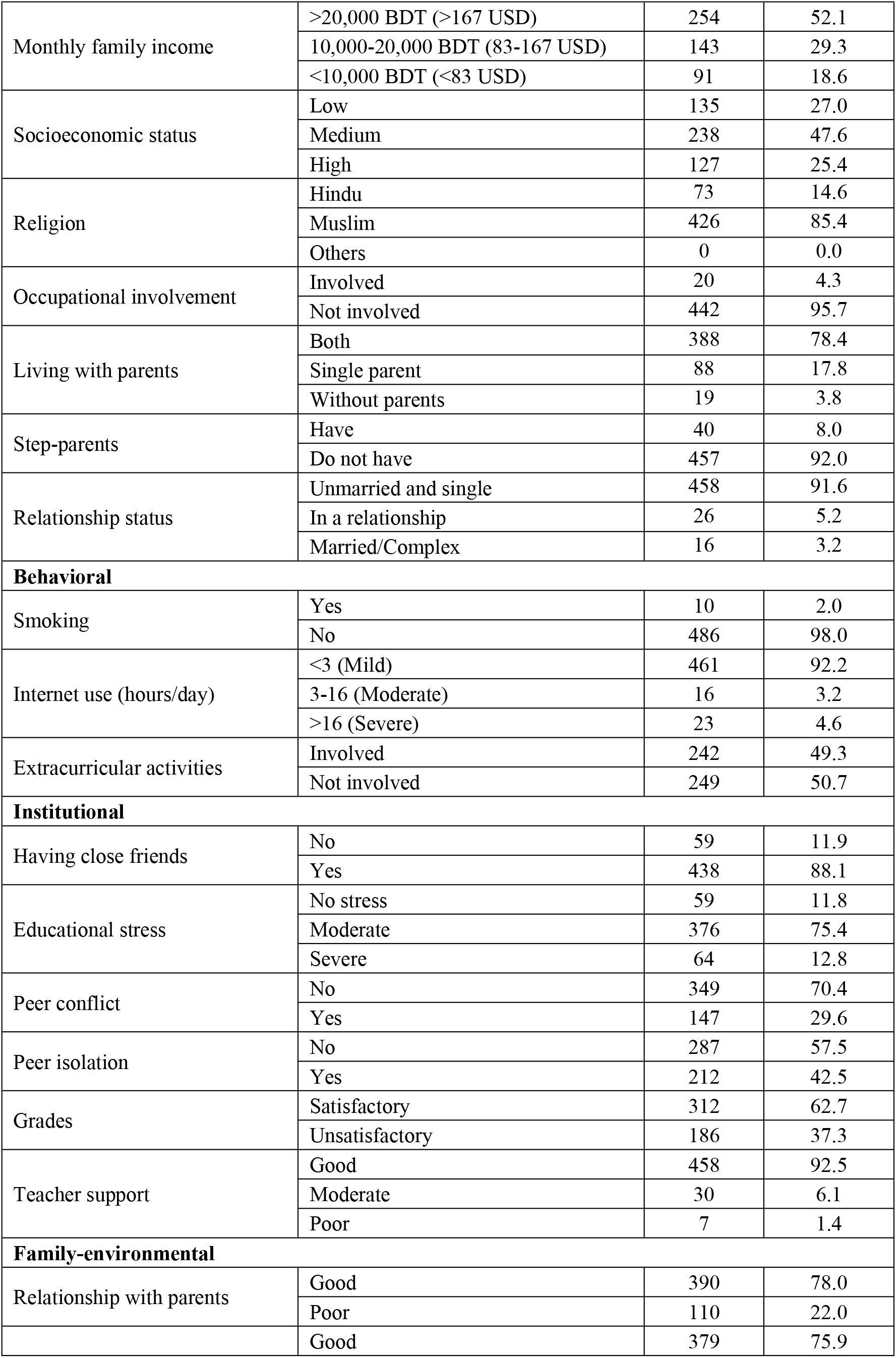
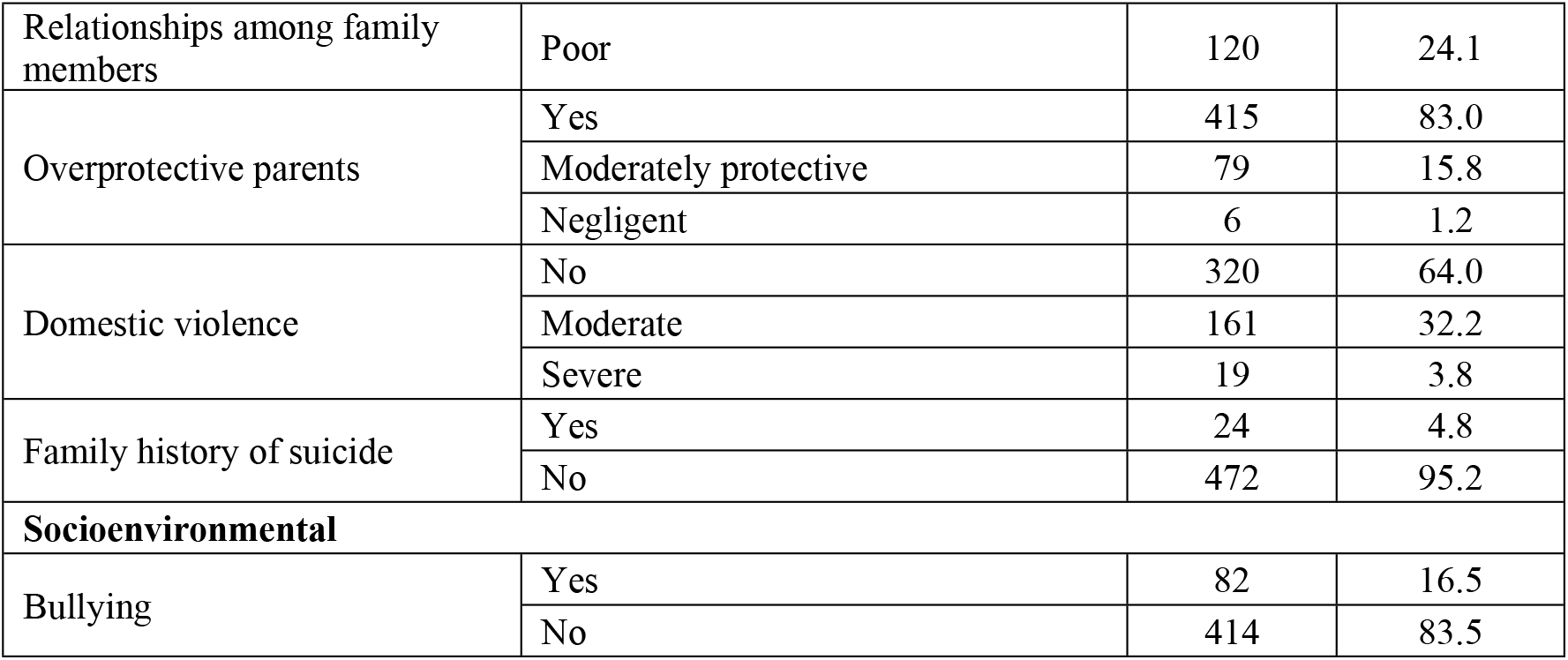
Baseline characteristics of adolescents aged 11–17 years (n = 500)

### Behavioral characteristics

Among the participants, 2.0% (n = 10) reported smoking, while no information was disclosed regarding substance use. Approximately 4.6% (n = 23) were classified as heavy internet users, spending more than 16 hours online per day [33]. Nearly half of the respondents (49.3%, n = 242) reported regular participation in extracurricular activities. **Table 1**.

### Institutional characteristics

Among the participants, 11.9% (n = 59) reported not having a close friend. Peer conflict was reported by 29.4% (n = 147), and 42.5% (n = 212) experienced peer isolation. Severe academic stress was reported by 12.8% (n = 64) of the participants, whereas 37.3% (n = 186) expressed dissatisfaction with their academic performance. The majority of the students (92.5%, n = 458) perceived their teachers as supportive. **Table 1**.

### Family-environmental characteristics

Approximately 22.0% (n = 110) of the participants reported having a strained relationship with their parents, whereas 24.1% (n = 120) indicated conflict among family members. Parental negligence regarding academic responsibilities was noted by 1.2% (n = 6) of the students. Severe domestic violence was reported by 3.8% (n = 19) of the respondents, and 4.8% (n = 24) reported a family history of suicide or suicide attempts.

### Socioenvironmental characteristics

Approximately 16.5% (n = 82) of the respondents reported experiencing bullying by classmates, teachers, family members, relatives, or neighbors. **Table 1**.

### Prevalence of stress symptoms among adolescents

As shown in **Table 2**, the overall prevalence of stress symptoms was 36.2% (n=181, 95% CI: 32.1%–40.5%). Mild stress symptoms were reported in 10.6% of the respondents (n = 53; 95% CI: 8.2%–13.6%), whereas 10.8% (n = 54; 95% CI: 8.4%–13.8%) experienced moderate stress symptoms. Severe stress symptoms were observed in 9.8% of the participants (n = 49; 95% CI: 7.5%–12.7%), and 5.0% (n = 25; 95% CI: 3.4%–7.3%) reported experiencing extremely severe stress symptoms.

**Table 2.** Prevalence of stress symptoms (n=500)

| Stress | DASS-21 score | Number (n) | Percentage (%) | 95% CI |
| --- | --- | --- | --- | --- |
| No stress | 0–14 | 319 | 63.8 | 59.5–67.9 |
| Mild | 15–18 | 53 | 10.6 | 8.2–13.6 |
| Moderate | 19–25 | 54 | 10.8 | 8.4–13.8 |
| Severe | 26–33 | 49 | 9.8 | 7.5–12.7 |
| Extremely severe | 34+ | 25 | 5.0 | 3.4–7.3 |

### Associated factors of stress symptoms

The chi-square test revealed several statistically significant associations between sociodemographic factors and stress levels (p < 0.05), as presented in **Table 3**. Specifically, age group 11–17 years (p < 0.001), gender (p = 0.001), socioeconomic status (p = 0.039), parental home ownership (p = 0.031), relationship status (p = 0.002), and living with step parents (p = 0.001) were significantly associated with stress symptoms.

**Table 3.**
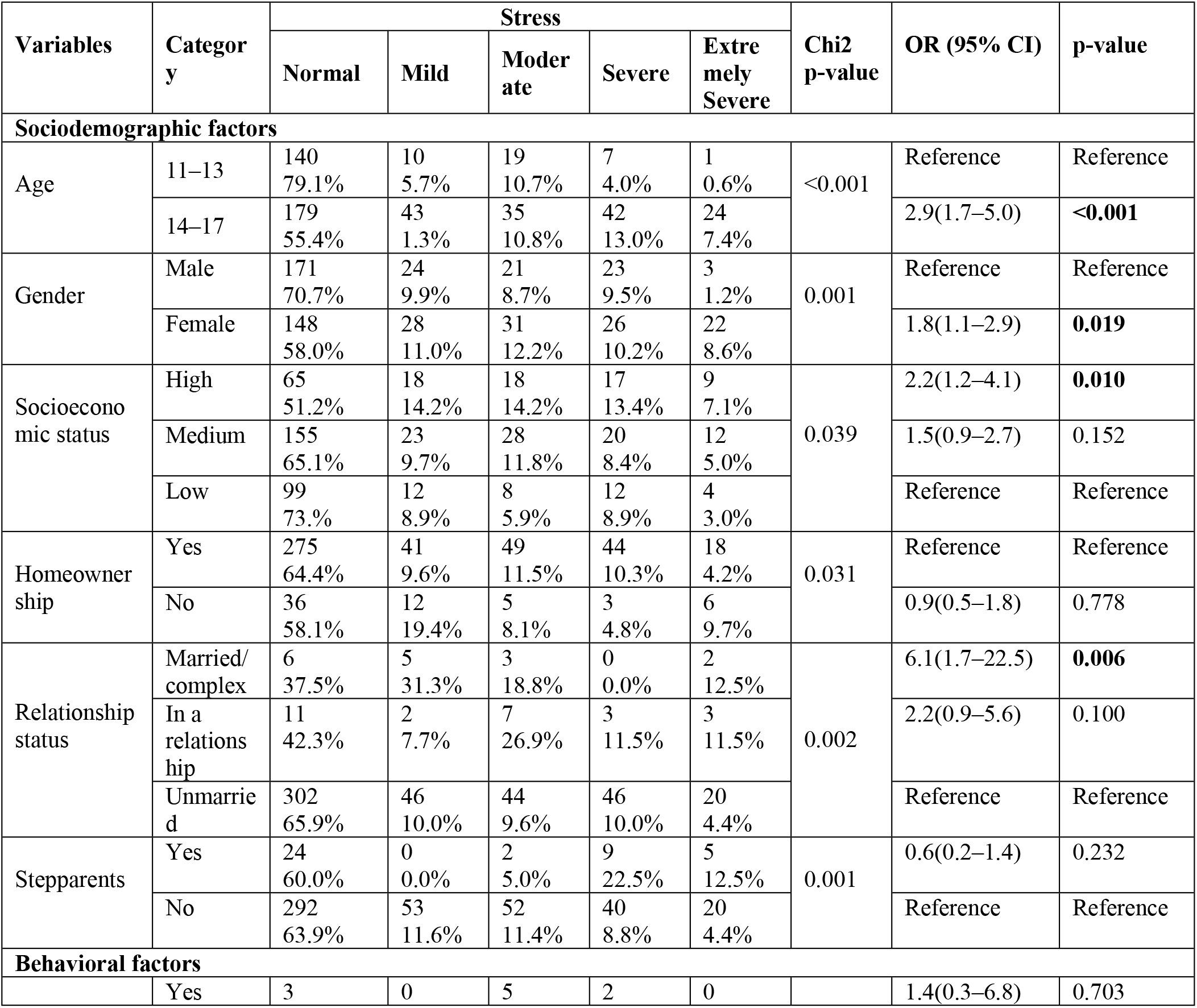
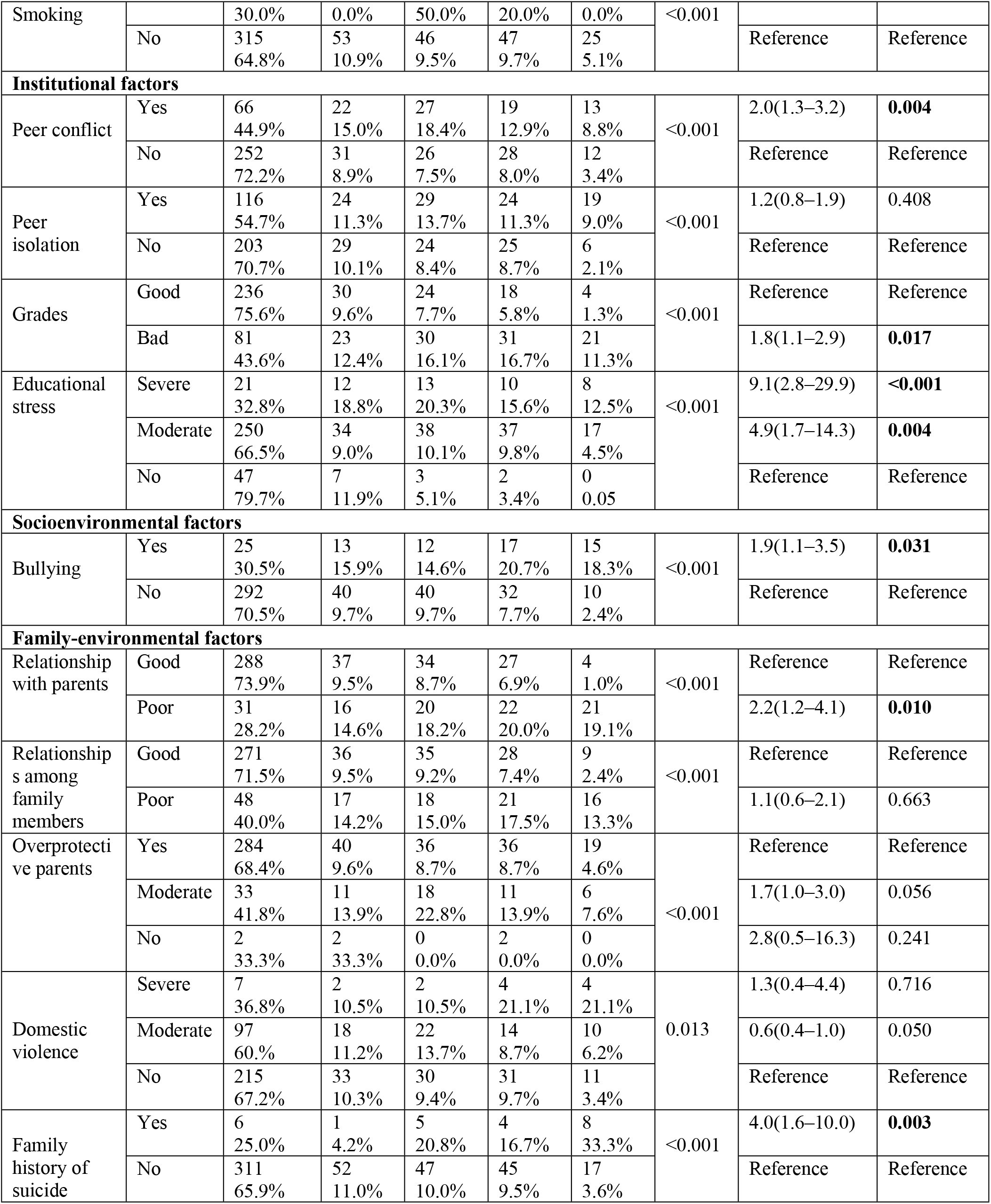
Factors associated with stress among adolescents (ordinal logistic regression)

Among the behavioral factors, smoking was strongly associated with stress symptoms (p < 0.001).

The significant institutional factors included peer conflict (p < 0.001), peer isolation (p < 0.001), academic performance (p < 0.001), and educational stress (p < 0.001).

In the domain of family-environmental factors, significant associations were observed for the quality of relationships with parents (p < 0.001), relationships with other family members (p < 0.001), parental overprotectiveness (p < 0.001), a family history of suicide (p < 0.001), and exposure to domestic violence (p = 0.013).

Additionally, being a victim of bullying emerged as a significant socioenvironmental factor related to stress symptoms (p < 0.001).

All variables identified as significant in the chi-square analysis were subsequently included in an ordinal logistic regression model to adjust for potential confounding factors. **Table 3**.

According to ordinal logistic regression analysis, adolescents aged 14–17 years presented nearly threefold increased odds of exhibiting stress symptoms (AOR: 2.9; 95% CI: 1.7–5.0; p < 0.001) compared with those aged 11–13 years. Compared with male adolescents, female adolescents were almost twice as likely to experience stress symptoms (AOR: 1.8; 95% CI: 1.1–2.9; p = 0.019). Similarly, those from high socioeconomic backgrounds had more than twice the odds of developing stress symptoms (AOR: 2.2; 95% CI: 1.2–4.1; p = 0.010) than their low socioeconomic counterparts. Compared with their unmarried peers, adolescents who were married or in complex relationships presented more than sixfold increased odds of stress symptoms (AOR: 6.1; 95% CI: 1.7–22.5; p = 0.006). Those experiencing severe educational stress had over ninefold increased odds of stress symptoms (AOR: 9.1; 95% CI: 2.8–29.9; p < 0.001), whereas those with moderate stress had nearly fivefold increased odds (AOR: 4.9; 95% CI: 1.7–14.3; p = 0.004) compared with those reporting no educational stress. Adolescents reporting peer conflicts had twice the odds of experiencing stress symptoms (AOR: 2.0; 95% CI: 1.3–3.2; p = 0.004), and those with poor academic performance had similarly elevated odds (AOR: 1.8; 95% CI: 1.1–2.9; p = 0.017).

Bullying victimization nearly doubled the odds of experiencing stress symptoms among adolescents (AOR: 1.9; 95% CI: 1.1–3.5; p = 0.031). Additionally, the likelihood of stress symptoms was two times greater (AOR: 2.2; 95% CI: 1.2–4.1; p = 0.010) in those with poor parental relationships and four times greater (AOR: 4.0; 95% CI: 1.6–10.0; p = 0.003) in those with a family history of suicide.

## Discussion

This study aimed to assess the prevalence and associated factors of stress symptoms among adolescents in rural Bangladesh. The prevalence of stress symptoms was 36.2% (95% CI: 32.1%–40.5%). Among the participants, 5.0% (95% CI: 3.4%–7.3%) experienced extremely severe stress symptoms, 9.8% (95% CI: 7.5%–12.7%) reported severe stress symptoms, 10.8% (95% CI: 8.4%–13.8%) experienced moderate stress symptoms, and 10.6% (95% CI: 8.2%– 13.6%) reported mild stress symptoms. Significant factors associated with stress symptoms included female sex, age 14–17 years, being married or in a complex relationship, higher socioeconomic status, moderate to severe academic stress, peer conflict, poor academic performance, experiences of bullying, strained parental relationships, and a family history of suicide.

A study conducted in four Southeast Asian countries (Malaysia, Indonesia, Singapore, and Thailand) reported a 36.0% prevalence of severe to extremely severe stress [34]. In Bangladesh, the prevalence of mental health disorders ranges from 6.5% to 31.0% among adults and from 13.4% to 22.9% among children [35]. A cross-sectional study in Chittagong, Bangladesh, using the Life Stress Scale highlighted that stress is more prevalent among urban adolescents than among their rural counterparts [25]. Similarly, a study conducted in Dhaka—the capital and urban setting—using the Perceived Stress Scale (PSS-10) reported that 65.0% of adolescents experienced moderate stress, and 9.0% experienced high stress [23].

Adolescence is the period when several mental health disorders emerge [3], particularly between the ages of 15–19, and is widely recognized as having a period of heightened stress levels due to academic pressure and social-emotional challenges [36]. Studies from Chennai, India [36] and various European countries [37] have confirmed increased stress levels during this stage, with adolescent females consistently reporting higher stress levels. A longitudinal study from Tennessee also demonstrated a progressive increase in stress symptoms from freshman to senior high school years, particularly among girls [38]. Our findings are consistent with these trends, showing elevated stress symptoms among adolescents aged 14–17, especially females.

High socioeconomic status does not necessarily shield adolescents from stress. Instead, it may introduce unique stressors such as academic pressure, parental expectations, and emotional disconnection. Multiple studies have shown that adolescents from affluent backgrounds are disproportionately affected by these factors [39-42], a pattern also observed in our study.

In Bangladesh, early marriage remains prevalent, with 78.2% of adolescents married before the age of 18 and 5.5% before the age of 13 [43]. Factors such as living with one’s in-laws, economic dependency, and family conflict contribute to psychological distress. Prior studies reported a 23.7% prevalence of stress among early-married girls in Bangladesh [44] and a 29.9% prevalence among partnered adolescents in Brazil [45]. Consistent with these findings, our study revealed a significant association between marital or complex relationship status and higher stress levels.

Academic and parent-related stressors are consistently identified as primary contributors to adolescent stress symptoms worldwide [46], which aligns with our findings. Strained parent−adolescent relationships are particularly impactful. A longitudinal study in China linked parental conflict with increased hopelessness, reduced life satisfaction, and general psychiatric morbidity [47]. Similarly, a Swedish cohort study revealed that adolescents reporting poor family relationships had an increased risk of psychiatric hospitalization later in life, independent of other adverse childhood experiences [48, 49]. Our study also identified familial conflict as a significant stressor.

Peer conflicts and academic challenges further exacerbate adolescent stress. Interpersonal issues among peers have been linked to emotional distress [50, 51], whereas poor academic performance and excessive expectations are associated with negative cognitive and emotional outcomes [52, 53]. Our results confirm that moderate and severe educational stress are among the leading contributors to adolescent stress symptoms.

Bullying is another critical factor. A meta-analysis of over 133,000 participants revealed that adolescents exposed to bullying were 2.8 times more likely to develop depression [54]. Bullying has also been associated with dysregulated physiological stress responses and increased risk for mental health disorders, indicating long-term impacts [55]. A review and meta-analysis highlighted a causal relationship between bullying victimization and various mental health issues [56].

A family history of suicide significantly increases the risk of adolescent stress. Adolescents with a parental history of suicide attempts are more likely to attempt suicide [57]. These individuals often experience greater emotional dysregulation [58] and a more stressful life context [59]. Similarly, our study identified a family history of suicide as a significant risk factor for adolescent stress symptoms.

## Limitations

As this is a cross-sectional study, causal inferences between variables cannot be established. This study utilized data collected from three selected schools within a short period due to time and resource constraints, which limits the ability to capture trends.

Although the admission policies of those institutions promote inclusivity, students from remote areas are often less likely to enroll due to geographic and logistical challenges. Furthermore, a significant number of adolescents residing in remote or underserved regions cannot access formal education because of financial, infrastructural, and sociocultural barriers. Consequently, the sample may not fully reflect the broader population, resulting in generalizability bias.

The categorization of internet usage was based on previously cited online sources. The validity of this threshold is not formally established, which may reduce precision in differentiating the usage levels.

Additionally, the data were based on self-reported responses, which may cause reporting bias or social desirability bias. Data on previous trauma, mental health disorders, and substance use were not collected, which could influence stress or potentially confound the results.

## Conclusion

Approximately three in ten adolescents in rural Bangladesh experience mild to extremely severe symptoms of stress. These findings underscore the urgent need for integrated, context-specific mental health interventions targeting adolescents in rural areas. Educationally and culturally appropriate programs, workshops, and competitions should be organized in schools to increase adolescent participation. They should be encouraged to engage in extracurricular and voluntary activities to reduce academic stress. The academic curriculum needs updating and organization, emphasizing cooperative learning rather than unhealthy competition. School authorities should prioritize the use of strict rules and regulations to create a healthy and supportive institutional environment.

Additionally, adolescents can be encouraged to participate in behavioral intervention programs at the community level. Parents and guardians should closely monitor children in this vulnerable age group and protect them from becoming involved in family conflicts. Additional psychological counseling and care are necessary for those with a family history of suicide.

Schools, families, and community organizations must collaborate to provide psychological support, reduce academic pressure, and foster supportive social environments. Further longitudinal and interventional studies are warranted to explore causality and inform effective, sustainable mental health strategies for this vulnerable population.

## Data Availability

Data is available in figshare DOI: https://doi.org/10.6084/m9.figshare.25854445.v4

## Acknowledgments

The authors express their sincere gratitude to the headmasters of the three participating schools for their invaluable support and cooperation during the data collection process.

## Supporting information

<u>S1 File.</u> Characteristics of adolescents aged 11–17 years in Lohagara, Narail, 2024.

DOI: https://doi.org/10.6084/m9.figshare.25854445.v4 [60]

